# Clinical validation of 3D-printed nasopharyngeal and oropharyngeal swabs for SARS-CoV-2 RT-PCR

**DOI:** 10.1101/2022.05.16.22274315

**Authors:** Åse Garløv Riis, Tonje Merethe Røssland, Iren Høyland Löhr, Ingvild Dalen, Lars Kåre Kleppe, Jon Sundal, Åse Berg, May Sissel Vadla, Ole Bernt Lenning, Heidi Syre

## Abstract

Due to limited access to commercially available flocked nasopharyngeal (NP) and oropharyngeal (OP) swabs during the SARS-COV-2 pandemic, we have evaluated the sensitivity of 3D-printed swabs compared to commercial swabs in a clinical setting. We included 35 subjects with known exposure to SARS-CoV-2. Participants were tested with commercial and prototype NP/OP swab pairs 8 and 22 days after exposure. At day 8, the sensitivity of the prototype was 96% for NP-samples (CI 81-99%) and 91% for OP-samples (CI 72-97%). The sensitivity of the commercial swab was 92% for NP-samples (CI 76-98%) and 91% for OP-samples (CI 72-97%). At day 22, the sensitivities of the commercial swab were 100% for NP-samples (CI 82-100%) and OP-samples (CI 77-100%), whereas sensitivity of the prototype was 61% for NP-samples (CI 39-80%) and 54% for OP-samples (CI 29-77%). In conclusion, the prototype might be an alternative to commercial swabs when used early in the course of infection.

## 1. Introduction

Coronavirus disease 2019 (COVID-19) is caused by severe acute respiratory syndrome coronavirus 2 (SARS-CoV-2) and was first described late 2019. The virus has spread widely throughout the world, and on March 11 2020 - the World Health Organization (WHO) declared a pandemic.

Large scale testing is a crucial step in controlling the ongoing COVID-19 pandemic, to identify infected individuals and for surveillance of novel virus variants [1].

National testing guidelines for COVID-19 varies between countries. The Infectious Disease Society of America (IDSA) recommends testing for all individuals with symptoms suspicious for COVID-19 [2].

One of the recommended methods for initial diagnostic testing for SARS-CoV-2 infection is nucleic acid amplification tests (NAAT) [3] from samples collected from an upper respiratory specimen [4]. Multiple Real Time reverse transcriptase (RT)-PCR assays targeting different genes of the SARS-CoV-2 genome have been developed during the course of the COVID-19 pandemic [5, 6].

WHO recommends using a method for detection of two different targets, including one target specific for COVID-19 or SARS-like coronavirus [7]. The cycle threshold (Ct) value obtained by RT-PCR represents the number of cycles needed in the PCR reaction to surpass the threshold for positive test results and is inversely related to the viral load [8]. Thus, the Ct value can potentially be used as an indirect estimate of the viral RNA copy number in a sample [8]. In a clinical setting SARS-CoV-2 RT-PCR test results are normally qualitatively reported as negative or positive without reporting the Ct value. There is no perfect standard for the evaluation of the clinical sensitivity of COVID-19 diagnostic tests. A meta-analysis, suggested that Nasopharyngeal (NP) swabs, or pooled nasal and oropharyngeal (OP) swabs, offer the best diagnostic performance among samples collected in the upper respiratory tract [9]. The clinical sensitivity may be influenced by several factors, including the prevalence in the population tested, the analytic sensitivity of the test assay, and the quality of the swab used. Hence, the false negative rates in samples collected from the nasopharynx varies from 1.8 to 33% in a systematic review, with an unexplained heterogeneity in the proportion of false negative RT-PCR results [10].

The increasing demand for SARS-CoV-2 testing during the pandemic placed pressure on supply chains. During the initial phase of the pandemic, limited access to diagnostic equipment, including commercially available flocked NP and OP swabs, was a well-known concern [11], 3D printing of NP and OP swabs could possibly reduce the problem. Callahan *et al* [12] evaluated 160 3D-printed NP swab designs from 24 different manufacturers. Four 3D-printed prototypes, including a prototype from HP inc, passed initial testing and completed a clinical trial. Reference and prototype swab pairs were collected from outpatients who came to a test station with COVID-19 suspicious symptoms. All four prototypes exhibited a high degree of concordance with the reference swab.

During a period of limited access to commercial flocked NP and OP swabs, and lack of 3D-printed swabs with CE approval in our region, we wanted to evaluate the utility and sensitivity of 3D-printed NP and OP swabs from a local producer. 3D-printed swabs were compared to commercial flocked NP and OP swabs used in our outpatient test stations. We included both symptomatic and asymptomatic patients with known exposure to SARS-CoV-2.

## 2. Materials and Methods

### 2.1. Subjects

Related to a bus tour for senior citizens in the South of Norway mid-September 2020, an outbreak of COVID-19 occurred. Seven days after departure, one of 40 passengers developed symptoms and tested positive for SARS-CoV-2, this was defined as day 0. The following seven days, the remaining 39 passengers underwent repeated testing with commercial OP, and for some cases with commercial NP swabs. In total 38 of 40 passengers tested positive for SARS-CoV-2. Two passengers remained PCR-negative. Thirty-five of the 40 passengers were included in the study. Participants gave informed consent.

Day 0-7 was defined as baseline. At baseline, the study population consisted of 33 PCR-positive and 2 PCR-negative individuals. The two PCR-negative individuals remained PCR-negative throughout the follow-up period, and did not develop SARS-CoV-2 specific antibodies. The proportion of asymptomatic, PCR-positive individuals was 24% (8/33), while 76% (25/33) developed symptomatic COVID-19.

### 2.2. Specimen collection

The 35 study participants underwent repeated testing with NP and OP swabs on day 8 - 10, and on day 22-24 (from now on referred to as day 8, and day 22). Commercial (FLOQSwabs, Copan, Italy) and prototype (3D-printed; HP inc, Norway) NP and OP swab pairs were collected from each participant in a random order. Each swab was placed in 3 ml sterile universal transport medium (UTM; Copan). Due to discomfort, some of the participants opposed repeated testing with NP swabs. An experienced general practitioner or infectious disease doctor performed sampling. The specimens were transported at room temperature to the laboratory and were stored at 4°C for up to 4 days prior to RT-PCR.

### 2.3. Laboratory analysis

All samples were analyzed at Stavanger University Hospital, Norway. NP and OP swabs, both commercial and prototypes, collected the same day for each patient, were processed and analyzed within the same batch for comparison. Viral RNA extraction was performed using the RNAdvance Viral Reagent Kit (Beckman Coulter; BC) and the Biomek i7 Automated Workstation (BC Life Sciences, Indianapolis, IN), according to the manufacturer’s instructions. The sample input volume was 200 µl while the nucleic acid elution volume was 40 µl. Each batch included two negative extraction controls consisting of UTM and an in-house prepared SARS-CoV-2-positive extraction control. RT-PCR was performed in a reaction mix of 5 µl sample eluate, 1x TaqMan Fast virus 1-step mix (Life Technologies), primers (0.5 µM) and probe (0.35 µM) from Tib-Molbiol targeting the *E*-gene of *Sarbecovirus* [7]. RT-PCR was performed on a QuantStudio 6 system (Applied Biosystems; AB, Waltham, MA, USA) with thermal cycling conditions of 50°C for 5 min., 95°C for 25 sec., 40 cycles at 95°C for 3 sec. and 60°C for 30 sec. QuantStudio Real-Time PCR Software (AB) was used for the presentation of the RT-PCR results. The analytical sensitivity and the limit of detection for the SARS-CoV-2 analysis was 20 RNA copies per reaction.

The results were categorized as negative, positive or weak positive based on interpretation of the Ct value and the amplification plot. Samples with increased fluorescence signal (ΔRn, normalized reporter value) below 0.05, were reported as negative. Samples with a Ct value equal to or below 35 were reported as positive, whereas samples with a Ct value above 35 or a ΔRn above 0.05 were reported as weak positive (low viral load), according to the laboratory clinical testing procedures. Due to a high pre-test probability (known outbreak in a defined group), the gold standard was defined as any positive or weak positive test result in either 3D printed or commercially available swabs for the respective sample sites and time points.

For data presentation, weak positive samples without a Ct value, but a ΔRn above 0.05 were assigned a Ct value of 40 (n=28; 16 commercial and 12 prototype) and negative samples were assigned a Ct value of 45 (n=110; 31 commercial and 79 prototype).

### 2.4. Statistical analysis

Diagnostic categories were compared between prototype swabs and commercial swabs by cross-tabulation. Chance-adjusted agreement was estimated as quadratically weighted Cohen’s Kappa (K) and Gwet’s AC2 coefficient and reported with t-distribution based 95% confidence intervals (CI). Gwet’s AC2 is often preferred in situations with uneven marginals, where Cohen’s kappa may give paradoxical results [13]. Differences between swabs were tested with McNemar’s test for paired data. Ct values are presented as medians and interquartile ranges (IQR), and compared between swab types for a given location and time point using Wilcoxon signed ranks tests.

The sensitivity of each swab type for a given sample site and time point was estimated by comparison with the gold standard, and presented with 95% Wilson CI. Because the study contains few test-negative individuals, specificity has not been calculated. Agreement statistics were estimated in R version 4.0 using functions ckap and gac from package rel; Wilson CIs and McNemar’s test for paired data were estimated in Stata v. 16.1; descriptive statistics and plots were created in SPSS v. 26. P-values <0.05 were considered statistically significant.

## 3. Results

We collected and analyzed 124 prototype and commercial swab pairs (60 NP and 64 OP) from 35 participants. Twenty-eight NP and 30 OP swab pairs were collected on day 8; remaining pairs were collected on day 22.

Figure 1 shows comparison of performances of NP and OP swabs at day 8 and day 22. Overall, when comparing performances of the two swab types with the gold standard, the prototype failed to detect 16 positive samples (15 weak positive), while the commercial swab failed to detect four (two weak positive).

**Figure 1:**
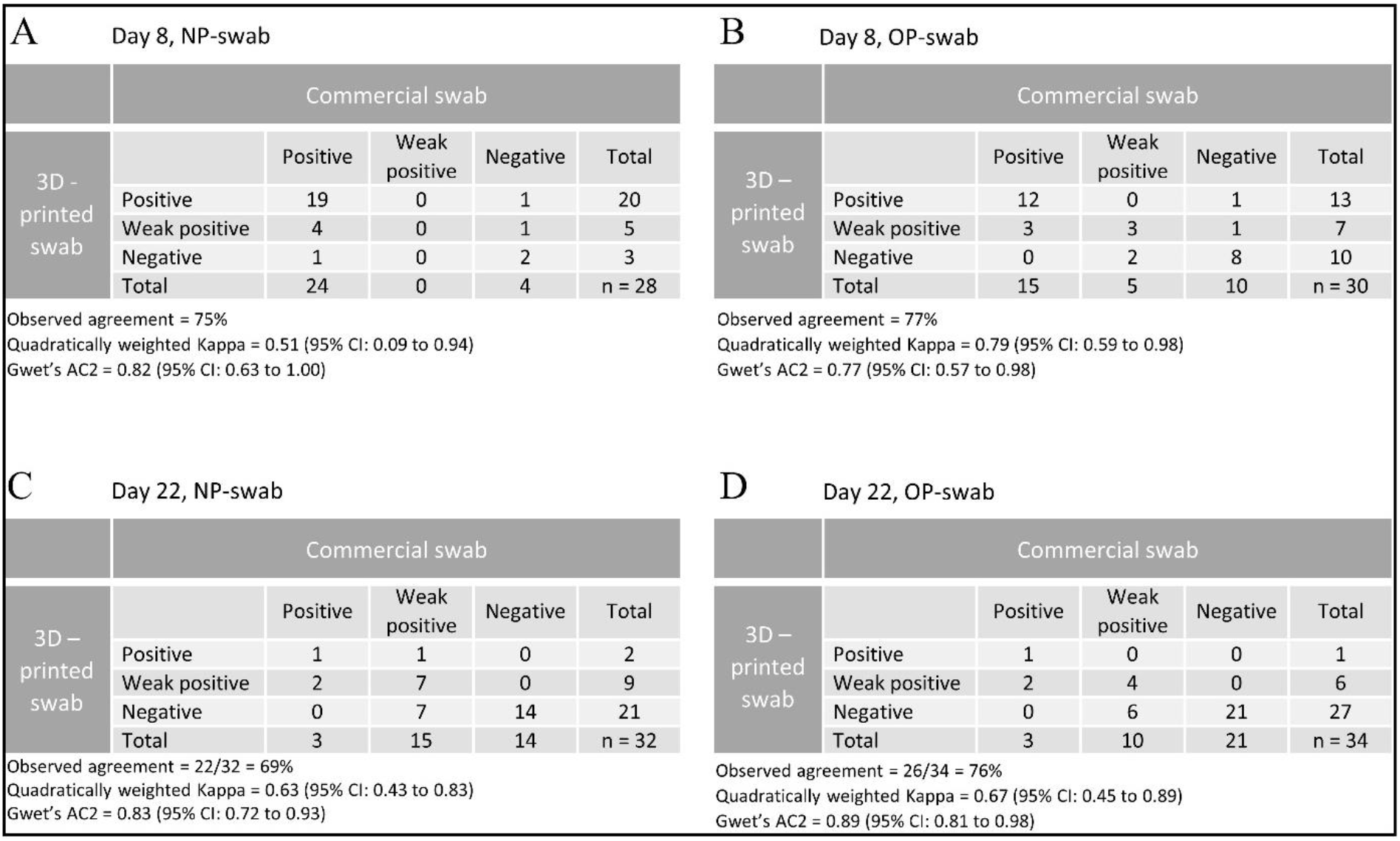
Agreement between 3D-printed swab and commercial swab (Copan) for day 8 and 22. Positive: Ct value ≤ 35. Weak positive: Ct value > 35 or ΔRn > 0.05. Negative: Ct value > 40 and ΔRn < 0.05. Abbreviations: Nasopharynx (NP) Oropharynx (OP). **A)** Classifications of NP samples collected with Commercial swab and 3D – printed swab at day 8 (n=28). **B)** Classifications of OP samples collected with Commercial swab and 3D – printed swab at day 8 (n=30). **C)** Classifications of NP samples collected with Commercial swab and 3D – printed swab at day 22 (n=32). **D:** Classifications of OP samples collected with Commercial swab and 3D – printed swab at day 22 (n=34).

Compared with gold standard at day 8, the prototype failed to detect one of 26 positive NP samples (sensitivity 96% CI 81-99%), and two of 22 positive OP samples (sensitivity 91% CI 72 - 97%). The commercial swab failed to detect two of 26 positive NP samples (sensitivity 92% CI 76-98%) and two of 22 positive OP samples (sensitivity 91% CI 72-97%). Agreement between the two swabs for day 8 are given in Figure 1, (A and B).

In contrast, isolated data from day 22 show a detection rate in favour of the commercial swab. The commercial swab detected all of the 18 positive NP samples (sensitivity 100% CI 82-100%) and the 13 positive OP samples (sensitivity 100%, CI 77-100%), while the prototype failed to detect seven of the 18 positive NP samples (sensitivity 61% CI 39-80%) and six of the 13 positive OP samples (sensitivity 54% CI 29-77%). Notably, all of the undetected samples were weak positives. Agreement between the two swabs for day 22 are given in Figure 1, (C and D).

Ct values were significantly higher for the prototype compared to commercial swabs at both day 8 and day 22 and for both OP and NP samples (p values from 0.002 to 0.017) (Table 1, Figure 2). Furthermore, Ct values were significantly higher at day 22 compared to day 8 (Table 2).

**Table 1:** Comparison of Ct values of samples collected with Commercial and 3D-printed swabs.

|  | # pairs | **Commercial swab**<br>Median (IQR) | **3D-printed swab**<br>Median (IQR) | *p* |
| --- | --- | --- | --- | --- |
| **NP Day 8** | 28 | 28 (25, 32) | 32 (28, 37) | 0.002 |
| **OP Day 8** | 30 | 35 (28, 45) | 38 (33, 45) | 0.008 |
| **NP Day 22** | 32 | 40 (38, 45) | 45 (39, 45) | 0.017 |
| **OP Day 22** | 34 | 45 (40, 45) | 45 (45, 45) | 0.004 |

NP Nasopharynx, OP Oropharynx, IQR Interquartile range (Q1, Q3). Weak positives without Ct value have been designated a value of 40. Negative samples have been designated a value of 45. P-values from Wilcoxon signed ranks tests.

**Table 2:**
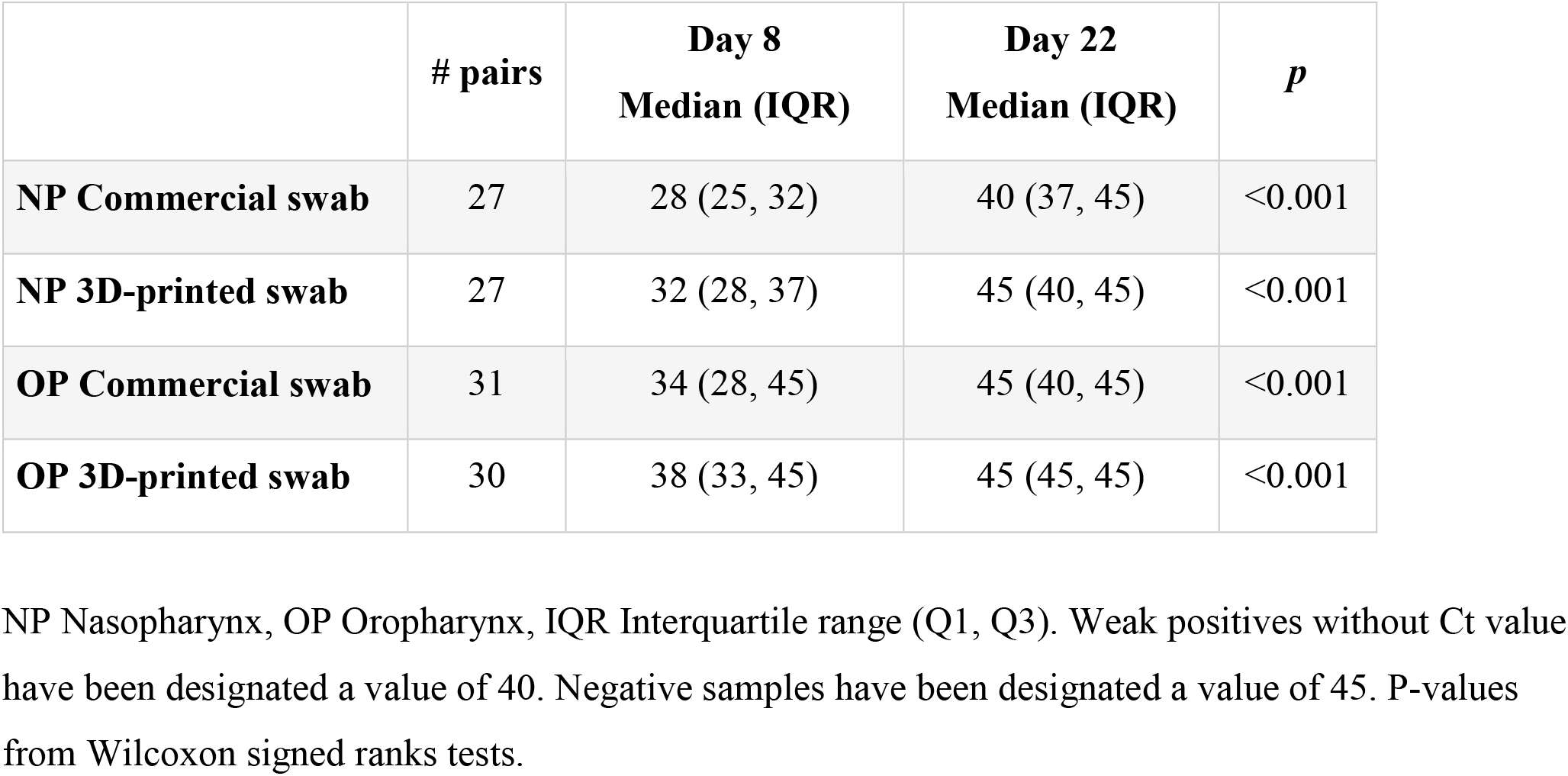
Comparison of Ct values of samples collected at day 8 and day 22

**Figure 2:**
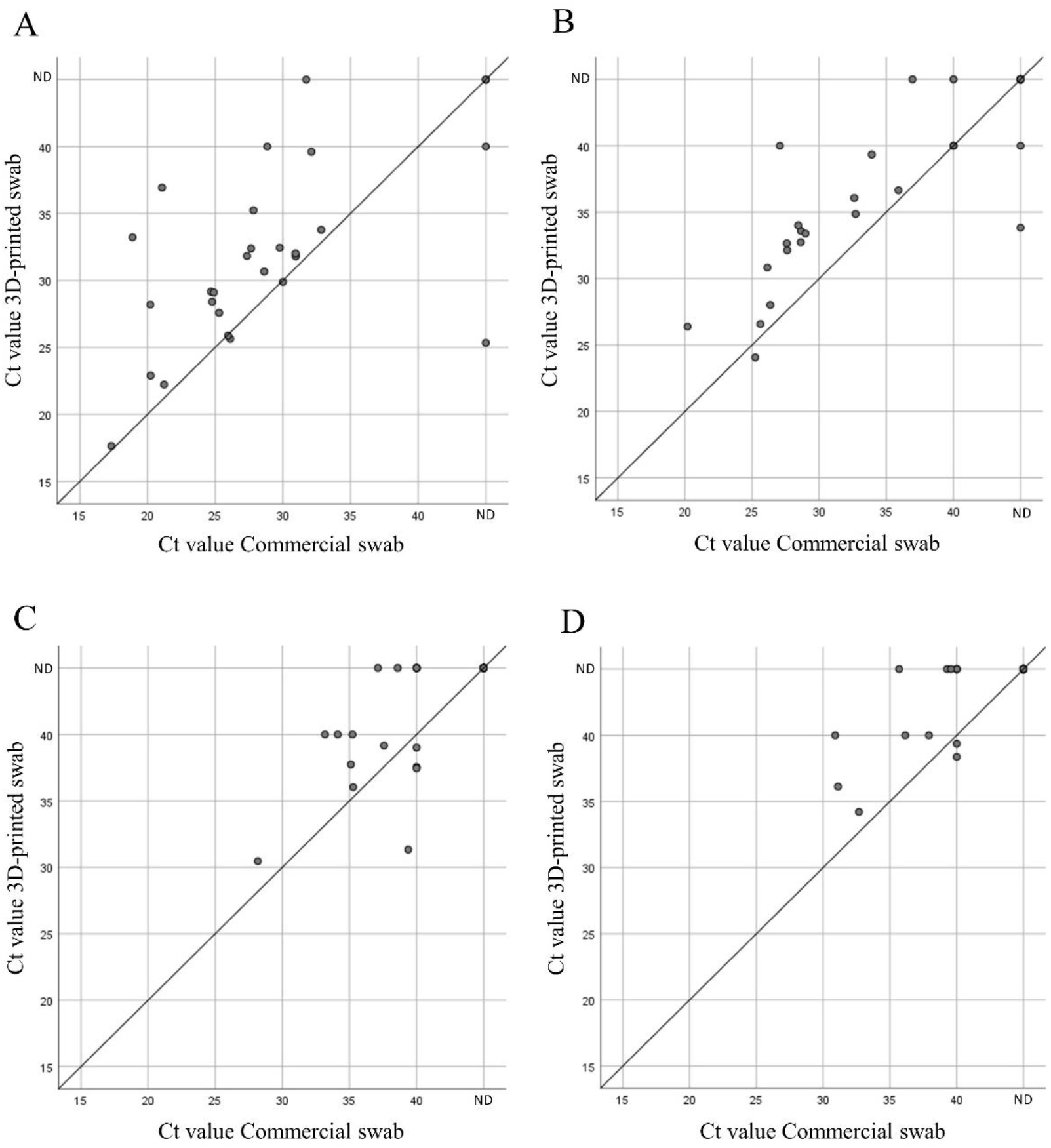
Comparison of Ct values for samples collected with 3D-printed prototype swab and samples collected with Copan commercial swab for **A)** Nasopharynx at day 8. **B)** Oropharynx at day 8. **C)** Nasopharynx at day 22. **D)** Oropharynx at day 22. Weak positives without Ct value have been designated a value of 40. Negative samples have been designated a value of 45, presented as not detected (ND). Lines of equality have been added to all plots. Each dot may represent several observations.

## 4. Discussion

The COVID-19 pandemic has resulted in severe shortages of medical diagnostic equipment; and in a period of limited access to commercially available flocked NP and OP swabs, we wanted to test the performance of a 3D-printed NP and OP swab for collection of samples for RT-PCR-analysis. This study shows that the 3D-printed prototype was non-inferior to the commercial swab (FLOQSwabs, Copan) in diagnosing infections with SARS-CoV-2 at day 8, and overall in samples with high viral loads. Ct values for the prototype were higher than for the commercial swab at day 8, but there was no difference in the rate of false negatives between the groups. At day 22, the prototype detected substantially less gold-standard positive samples, than the commercial swab. Most samples had low viral loads at day 22.

The results from day 8 are consistent with a previous clinical trial of outpatients with COVID-19 suspicious symptoms [12]. But in contrast to Callahan *et al*. [12] we found significantly higher Ct values for the prototype compared to the commercial swab, for all samples, including those collected at day 8, i.e. shortly after exposure to SARS-CoV-2 (P-values from 0.002 to 0.017). In patients infected with SARS-CoV-2, viral loads are reported to peak at symptom onset or during the first week of illness [14, 15], followed by a gradual decline [14-16]. Mean duration of SARS-CoV-2 RNA shedding from the upper respiratory tract is reported to be around 17 days in patients with mild to moderate illness [16, 17], and 19.8 days in severely ill patients [17].

According to a systematic review, live virus from respiratory samples could not be isolated beyond nine days of illness [16]. Isolation of live virus from respiratory specimens has been described beyond this period, and is probably more common in patients with severe illness [17, 18]. A sensitive test during the period where the viral load is at highest is critical to reduce further transmission of the virus. Nevertheless, there is potential harm in a false negative test, also later in the course of infection. This could affect COVID-19 public health management strategies in terms of case identification and contact tracing [19].

We collected the first swab-pairs at day 8, and have not evaluated the sensitivity of the prototype the first days after SARS-CoV-2 exposure. However, viral loads in the upper respiratory tract are reported to gradually increase towards symptom onset [16]. A study from 2020 suggest that the false negative rate of RT-PCR based tests decreases from time of exposure, and is lowest 8 days after exposure [20]. Thus, we consider the timing of testing at day 8 appropriate for comparing the performance of the swabs at the expected peak of viral load. As for day 22, it is possible that the prototype would under-perform compared to the commercial swab also during the first days after SARS-CoV-2 exposure and/or before symptom onset, when the viral load is expected to be low.

The proportion of asymptomatic individuals in our study was 24%. Viral loads in samples collected from the upper respiratory tract inferred from Ct values, are reported to be similar [21, 22] or lower [23] in asymptomatic cases. Furthermore, for asymptomatic cases, the period of viral shedding is reported to be shorter [24, 25], whereas in other studies statistically significant differences were not found [21, 23].

This study has some limitations. Testing was conducted on a low number of participants, and almost all the participants were SARS-CoV-2 positive, thus the study was not suitable for evaluating the specificity of the prototype. Furthermore, the usability of the prototype was not assessed. Thus, further testing is needed to determine the test-accuracy of 3D-printed swabs.

One strength of the study is that we repeated testing with both swab types shortly after exposure and later in the course of infection; this enabled us to evaluate the performance of the prototype on samples with different viral loads. Because our study population consisted of a defined group with known exposure to SARS-CoV-2, we were able to include both symptomatic and asymptomatic individuals. Finally, sample collection techniques can affect the clinical sensitivity of the test. In our study, experienced personnel performed sampling, which reduced the risk of false negative results. To prevent swab shortages during future pandemics, or seasonal epidemics, caused by influenza or other viruses, 3D printing of swabs might help to maintain rapid, large scale diagnostic testing.

## 5. Conclusion

In summary, this study shows that the performance of the 3D-printed prototype was non-inferior to the commercial swab eight days after exposure to SARS-CoV-2. In a shortage situation, the prototype might be an alternative to a commercial swab when used early in the course of infection or within eight days after known exposure.

## Data Availability

All data produced in the present study are available upon reasonable request to the authors.

## Abbreviations

OP: Oropharyngeal
NP: Nasopharyngeal
SARS-CoV-2: Severe Acute Respiratory Syndrome Coronavirus 2
WHO: World Health Organization
IDSA: Infectious Disease Society of America
COVID-19: Coronavirus disease 2019
CDC: Centers for Disease Control and Prevention
Ct: Cycle threshold

## Author contribution

The authors confirm contribution to the paper as follows: study conception and design: IHL, HS; data collection: ÅGR, TMR, JS, ÅB, MSV, OBL, HS; analysis and interpretation of results: ÅGR, TMR, ID, IHL, HS; draft manuscript preparation: ÅGR, TMR. All authors reviewed the results and approved the final version of the manuscript.

## Acknowledgements

We want to thank Hans Petter Torvik for recruiting participants, and contributing in conduction of the clinical trial. Jorunn Nilsen, Sanna Palmqvist, Siri Øksnevad, and Linda Gloppen for their assistance in collection of patient samples. Hanne Røland Hagland who introduced us to the manufacturer of the 3D-printed swabs.

## Ethical approval

The Regional Committee for Medical Research Ethics Western Norway (REK West) gave ethical approval for this work (reference no. 118664).

## References

1. Vogels, C.B.F., et al., Multiplex qPCR discriminates variants of concern to enhance global surveillance of SARS-CoV-2. PLOS Biology, 2021. 19(5): p. e3001236.

2. Hanson, K.E., et al., Infectious Diseases Society of America Guidelines on the Diagnosis of COVID-19. Clin Infect Dis, 2020.

3. CDC. Overview of Testing for SARS-CoV-2, the virus that causes COVID-19. Updated Feb. 11, 2022 [cited 2022 27.04].

4. CDC. Interim Guidelines for Collecting and Handling of Clinical Specimens for COVID-19 Testing. Updated Oct. 25, 2021 [cited 2022 19.04].

5. Mathuria, J.P., R. Yadav, and Rajkumar, Laboratory diagnosis of SARS-CoV-2 - A review of current methods. Journal of Infection and Public Health, 2020. 13(7): p. 901–905.

6. Asrani, P., et al., Diagnostic approaches in COVID-19: clinical updates. Expert Rev Respir Med, 2021. 15(2): p. 197–212.

7. Corman, V.M., et al., Detection of 2019 novel coronavirus (2019-nCoV) by real-time RT-PCR. Euro surveillance : bulletin Europeen sur les maladies transmissibles = European communicable disease bulletin, 2020. 25(3): p. 2000045.

8. Rao, S.N., et al., A Systematic Review of the Clinical Utility of Cycle Threshold Values in the Context of COVID-19. Infect Dis Ther, 2020. 9(3): p. 573–586.

9. Tsang, N.N.Y., et al., Diagnostic performance of different sampling approaches for SARS-CoV-2 RT-PCR testing: a systematic review and meta-analysis. The Lancet Infectious Diseases.

10. Arevalo-Rodriguez, I., et al., FALSE-NEGATIVE RESULTS OF INITIAL RT-PCR ASSAYS FOR COVID-19: A SYSTEMATIC REVIEW. PLOS ONE, 2020: p. 2020.04.16.20066787.

11. Vermeiren, C., et al., Comparison of Copan ESwab and FLOQSwab for COVID-19 Diagnosis: Working around a Supply Shortage. Journal of clinical microbiology, 2020. 58(6): p. e00669–20.

12. Callahan, C.J., et al., Open Development and Clinical Validation of Multiple 3D-Printed Sample-Collection Swabs: Rapid Resolution of a Critical COVID-19 Testing Bottleneck. Journal of clinical microbiology, 2020.

13. Quarfoot, D. and R.A. Levine, How Robust Are Multirater Interrater Reliability Indices to Changes in Frequency Distribution? The American Statistician, 2016. 70(4): p. 373–384.

14. He, X., et al., Temporal dynamics in viral shedding and transmissibility of COVID-19. Nat Med, 2020. 26(5): p. 672–675.

15. Wölfel, R., et al., Virological assessment of hospitalized patients with COVID-2019. Nature, 2020. 581(7809): p. 465–469.

16. Cevik, M., SARS-CoV-2, SARS-CoV, and MERS-CoV viral load dynamics, duration of viral shedding, and infectiousness: a systematic review and meta-analysis. Lancet microbe, 2020.

17. Fontana, L.M., et al., Understanding viral shedding of severe acute respiratory coronavirus virus 2 (SARS-CoV-2): Review of current literature. Infection control and hospital epidemiology, 2020: p. 1–10.

18. Kim, M.-C., et al., Duration of Culturable SARS-CoV-2 in Hospitalized Patients with Covid-19. New England Journal of Medicine, 2021. 384(7): p. 671–673.

19. Organization, W.H., Considerations for quarantine of contacts of COVID-19 cases: interim guidance, 19 August 2020. World Health Organization.. 2020.

20. Kucirka, L.M., et al., Variation in False-Negative Rate of Reverse Transcriptase Polymerase Chain Reaction-Based SARS-CoV-2 Tests by Time Since Exposure. Ann Intern Med, 2020. 173(4): p. 262–267.

21. Lee, S., et al., Clinical Course and Molecular Viral Shedding Among Asymptomatic and Symptomatic Patients With SARS-CoV-2 Infection in a Community Treatment Center in the Republic of Korea. JAMA Internal Medicine, 2020. 180(11): p. 1447–1452.

22. Ra, S.H., et al., Upper respiratory viral load in asymptomatic individuals and mildly symptomatic patients with SARS-CoV-2 infection. Thorax, 2021. 76(1): p. 61–63.

23. Zhou, R., et al., Viral dynamics in asymptomatic patients with COVID-19. Int J Infect Dis, 2020. 96: p. 288–290.

24. Noh, J.Y., et al., Asymptomatic infection and atypical manifestations of COVID-19: Comparison of viral shedding duration. The Journal of infection, 2020. 81(5): p. 816–846.

25. Yang, R., X. Gui, and Y. Xiong, Comparison of Clinical Characteristics of Patients with Asymptomatic vs Symptomatic Coronavirus Disease 2019 in Wuhan, China. JAMA Netw Open, 2020. 3(5): p. e2010182.

